# Unsupervised Clustering Applied to Electronic Health Record-derived Phenotypes in Patients with Heart Failure

**DOI:** 10.1101/2022.10.31.22281772

**Authors:** Nosheen Reza, Yifan Yang, William P. Bone, Pankhuri Singhal, Anurag Verma, Srinivas Denduluri, Srinath Adusumalli, Marylyn D. Ritchie, Thomas P. Cappola

## Abstract

**Background:** High-dimensional electronic health records (EHR) data can be used to phenotype complex diseases. The aim of this study is to apply unsupervised clustering to EHR-based traits derived in a cohort of patients with heart failure (HF) from a large integrated health system.

**Methods:** Using the institutional EHR, we identified 8569 patients with HF and extracted 1263 EHR-based input features, including clinical, echocardiographic, and comorbidity data, prior to the time of HF diagnosis. Principal component analysis, Uniform Manifold Approximation and Projection, and spectral clustering were applied to the input features after sex stratification of the cohort. The optimal number of clusters for each sex-stratified group was selected by highest Silhouette score and by within-cluster and between-cluster sums of squares. Determinants of cluster assignment were evaluated.

**Results:** We identified four clusters in each of the female-only (44%) and male-only (56%) cohorts. Sex-specific cohorts differed significantly by age of HF diagnosis, left ventricular chamber size, markers of renal and hepatic function, and comorbidity burden (all p<0.001). Left ventricular ejection fraction was not a strong driver of cluster assignment.

**Conclusion:** Readily available EHR data collected in the course of routine care can be leveraged to accurately classify patients into major phenotypic HF subtypes using data driven approaches.

## INTRODUCTION

The classification of heart failure (HF) as a clinical syndrome is imprecise. Historically, HF has been partitioned and studied through use of clinically convenient measurements, such as left ventricular ejection fraction (LVEF) and New York Heart Association (NYHA) functional classification. These classifications – used as tools for risk stratification, determination of clinical trial eligibility, and candidacy for drugs and devices – fail to capture etiologic and pathophysiologic heterogeneity.^1^ Specific subpopulations of patients with HF manifest disease and treatment response in distinct ways, supporting the need to refine the HF phenotype as an essential step to improve diagnosis, treatment, and outcomes.^2^

The use of high dimensional phenotypic data offers a new approach to identify subtypes of complex disease syndromes, like HF; however, how best to consolidate these data into clinically relevant and actionable information has been a challenge. Modeling approaches using unsupervised machine learning have successfully incorporated dense, high dimensional data for phenotype discovery in disease states like chronic obstructive pulmonary disease^3,4^ and type 2 diabetes.^5^ Clustering analysis, a type of hypothesis-free unsupervised machine learning which can classify subjects into groups based on inter-individual similarity, has been applied to well-characterized cohorts of patients with HF derived from clinical trial participants^6^ and national registries.^7^ This technique has yielded subtypes of chronic HF that differed significantly in outcomes and in response to therapeutics and that were otherwise unidentified using traditional phenotypic distinctions like LVEF.

The majority of machine learning approaches applied to groups of patients with HF have been derived from small, highly curated and comprehensively annotated datasets, like those from clinical trials.^6,8^ Such studies have shown that cluster analysis is able to uncover discrete phenotypic groups of patients with HF. A natural next step is to evaluate whether similar data-driven approaches based on large collections of real-world quantitative and qualitative features collected in routine clinical care, such as those that can be extracted from longitudinal electronic health records (EHR), can be integrated to define distinct HF subgroups and potentially uncover unrecognized drivers of differentiation into these subgroups. In this proof-of-concept analysis, we applied unsupervised clustering to EHR-based traits derived in a cohort of patients with HF from the Penn Medicine health system. We hypothesized that through deep EHR-based phenotyping, distinct phenotypic HF subgroups would be able to be identified.

## METHODS

### EHR Data Collection

We used the Penn Medicine EHR reporting database (Clarity, Epic Systems Inc., Verona, Wisconsin) to identify all patients who had an International Classification of Diseases, Tenth Revision, Clinical Modification (ICD-10-CM) diagnosis of HF from October 1998 to October 2019. HF was defined by the presence of a standard EHR grouper (grouper ID 100505) of diagnosis codes for HF listed in the active EHR problem list, medical history, or as a prior encounter (hospital, emergency department, office visit) diagnosis for at least 2 unique encounters. Date of HF diagnosis was designated as the date of first qualifying HF encounter. Those patients who did not have an echocardiogram within 5 years prior to HF diagnosis were excluded.

### Model Input Features

A total of 1263 features were collected from the EHR (Figure 1). The twenty-six quantitative clinical features included age at HF diagnosis, sex, smoking status, height, weight, systolic and diastolic blood pressures, heart rate, sodium, potassium, blood urea nitrogen, creatinine, estimated glomerular filtration rate, alanine aminotransferase, aspartate aminotransferase, alkaline phosphatase, total bilirubin, hemoglobin, total cholesterol, high density lipoprotein cholesterol, low density lipoprotein cholesterol, triglycerides; four echocardiographic measurements: LVEF, left ventricular posterior wall thickness, interventricular septal wall thickness, and left ventricular internal diameter in diastole; and 1235 ICD-10-CM codes. Quantitative (continuous variables) (i.e., laboratory values, vital signs, echocardiographic measurements) were extracted at the time closest prior to HF diagnosis.

**FIGURE 1.**
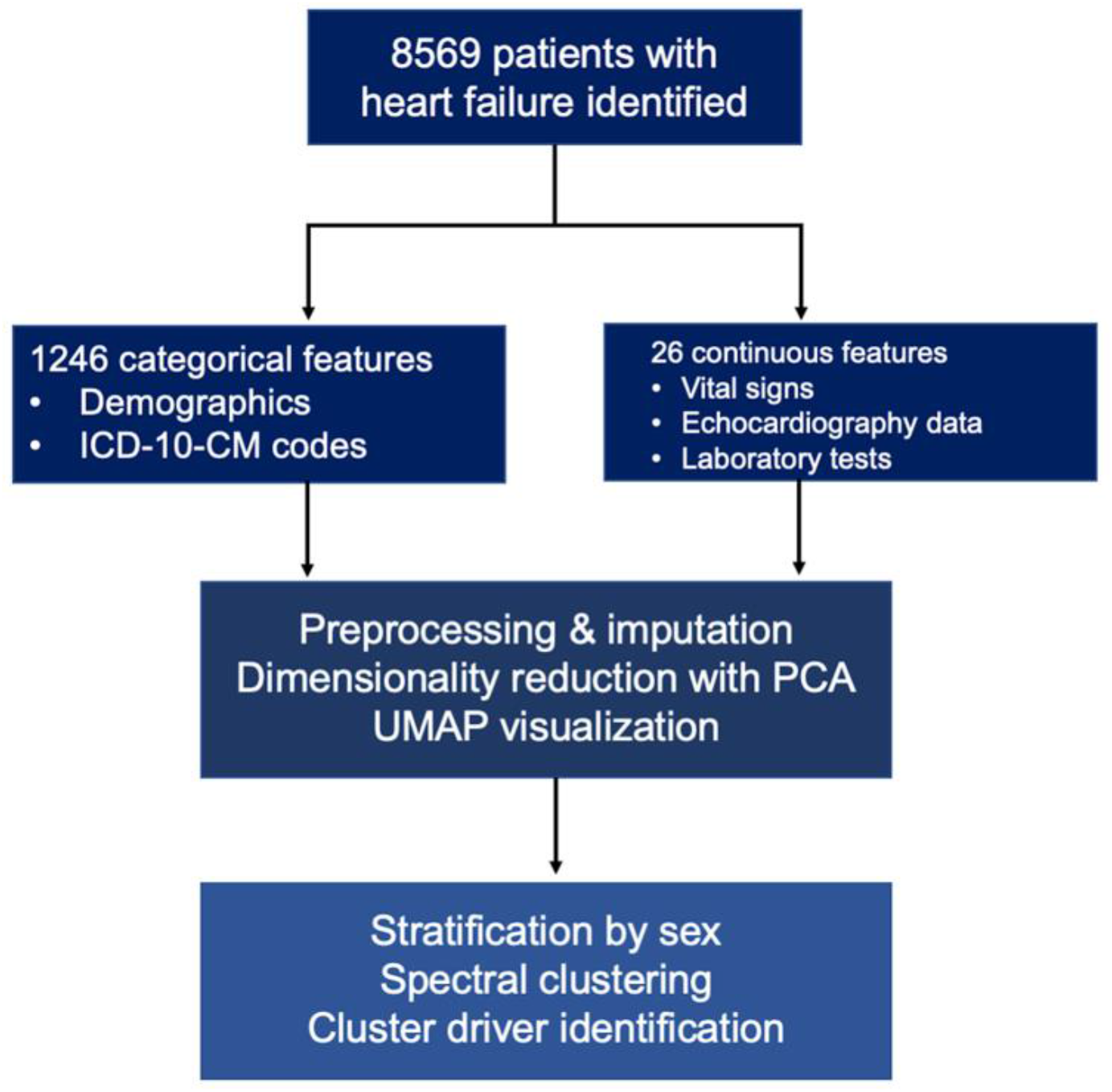
Study design.

### Data Preprocessing and Imputation

Patients with more than 10% missingness in their features were excluded, leaving a cohort with 90% or more completeness across features. Residual missing values were then imputed using the *missForest* R package, a nonparametric imputation method using random forest (normalized mean squared error representing error derived from imputing continuous values was 0.000059%). After quality control, the analytic cohort consisted of 8569 unique patients each with 1263 features.

### Dimensionality Reduction and Visualization

To represent these high-dimensional data into lower dimensional space, Principal Components Analysis (PCA) was performed, which yielded 543 principal components that explained 90% of the variance. The principal components were then entered into Uniform Manifold Approximation and Projection (UMAP) application to create a two-dimensional representation of the data. As sex was a disproportionately strong driver of dimension reduction, the cohort was stratified by female (N=3,800) versus male (N=4,769) sex (Figures 2 and 3).

**FIGURE 2.**
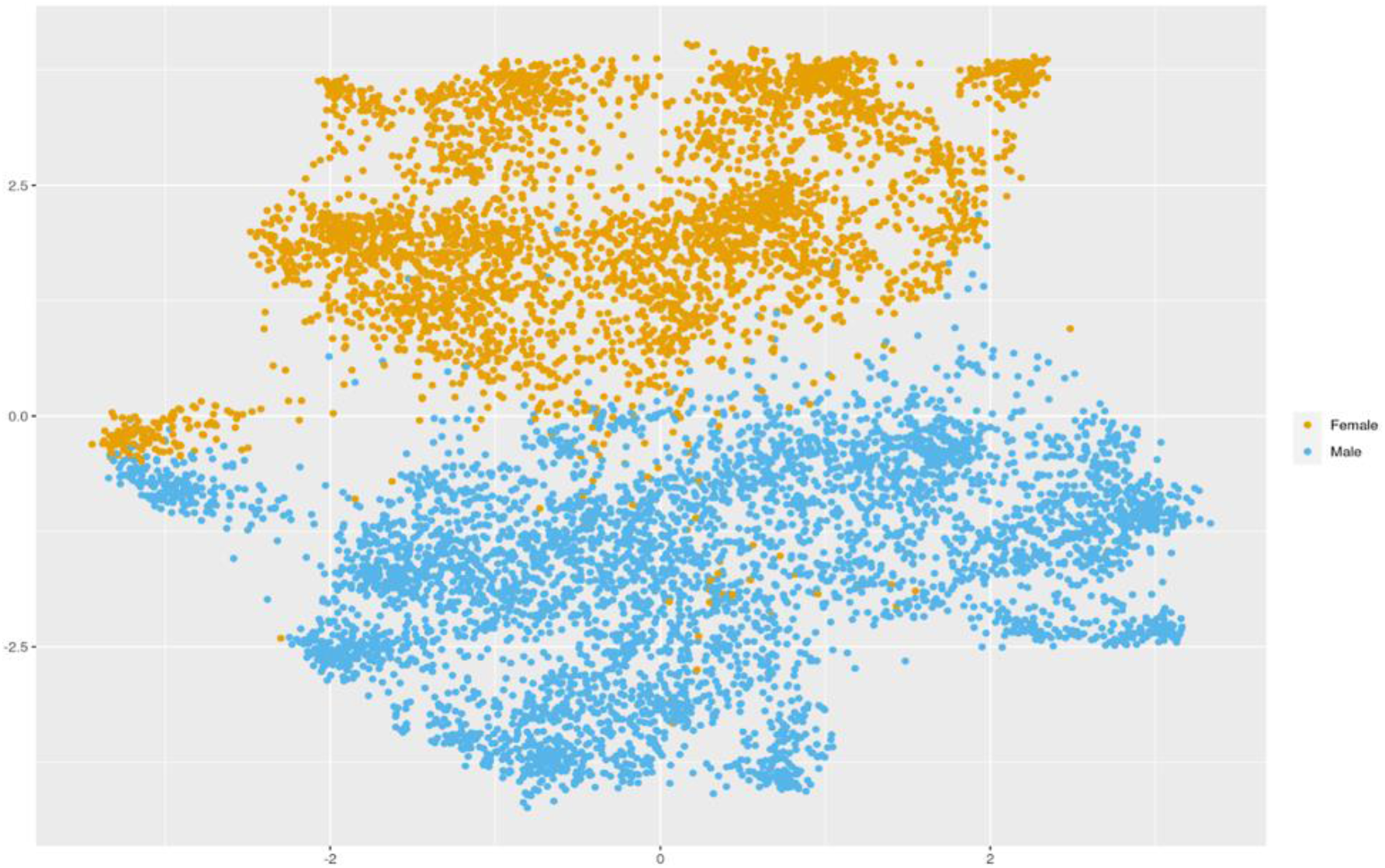
Uniform Manifold Approximation and Projection (UMAP) visualization of the patient cohort.

**FIGURE 3.**
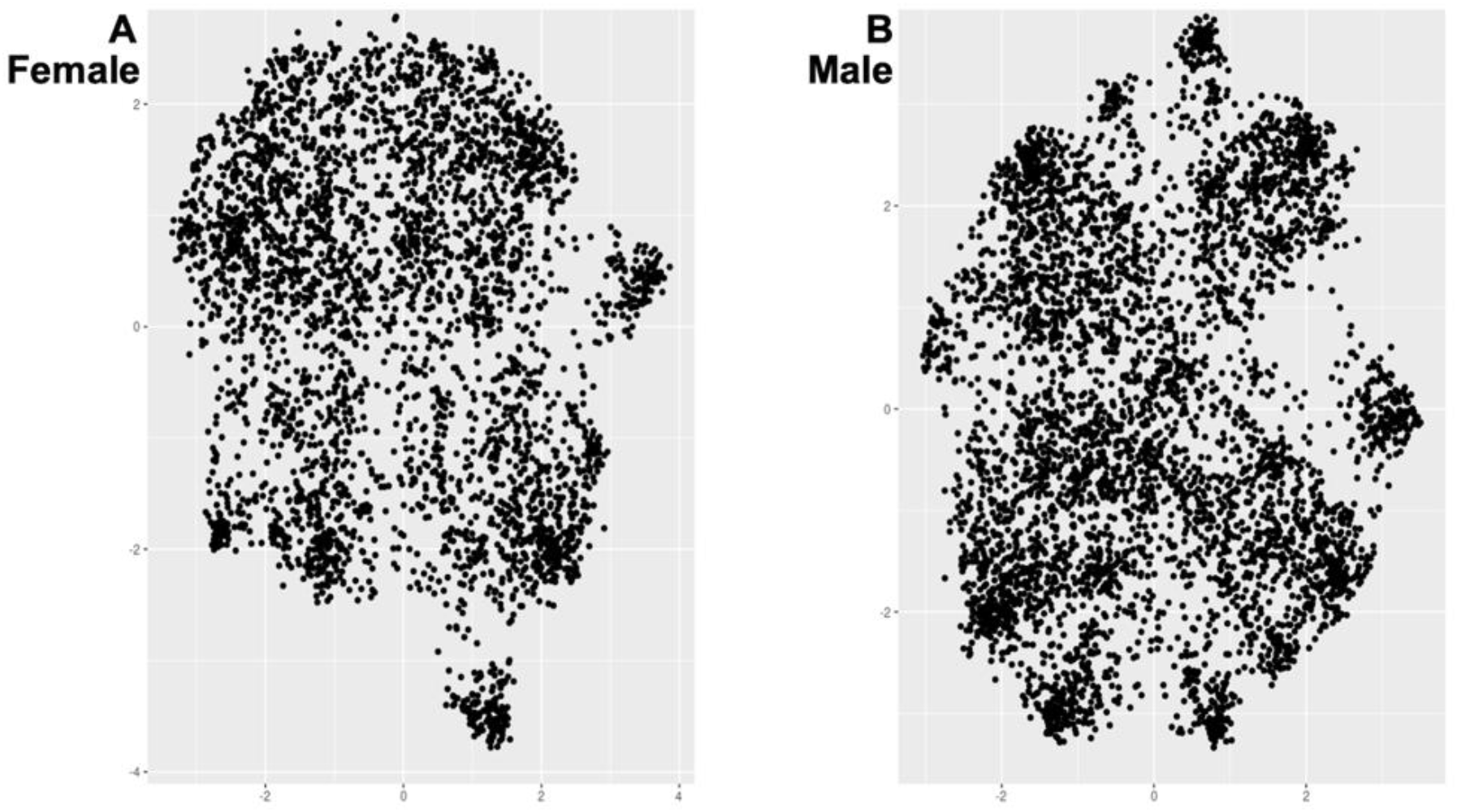
Uniform Manifold Approximation and Projection (UMAP) visualization of the patient cohort stratified by sex, (A) female and (B) male.

### Clustering Analysis

Spectral clustering on the UMAP representation was applied to each sex-stratified group. The optimal number of clusters for each sex-stratified group was selected by highest Silhouette score and by the maximum within-cluster and between-cluster sums of squares. For both females (Figure 4A) and males (Figure 4B), the optimal number of clusters was four. The vtest^9^ was performed to compare the proportion of EHR-derived traits in each cluster with the proportion of the EHR-derived traits in the study population. The sign of the test value indicates whether the trait is over- or under-represented in the cluster.

**FIGURE 4.**
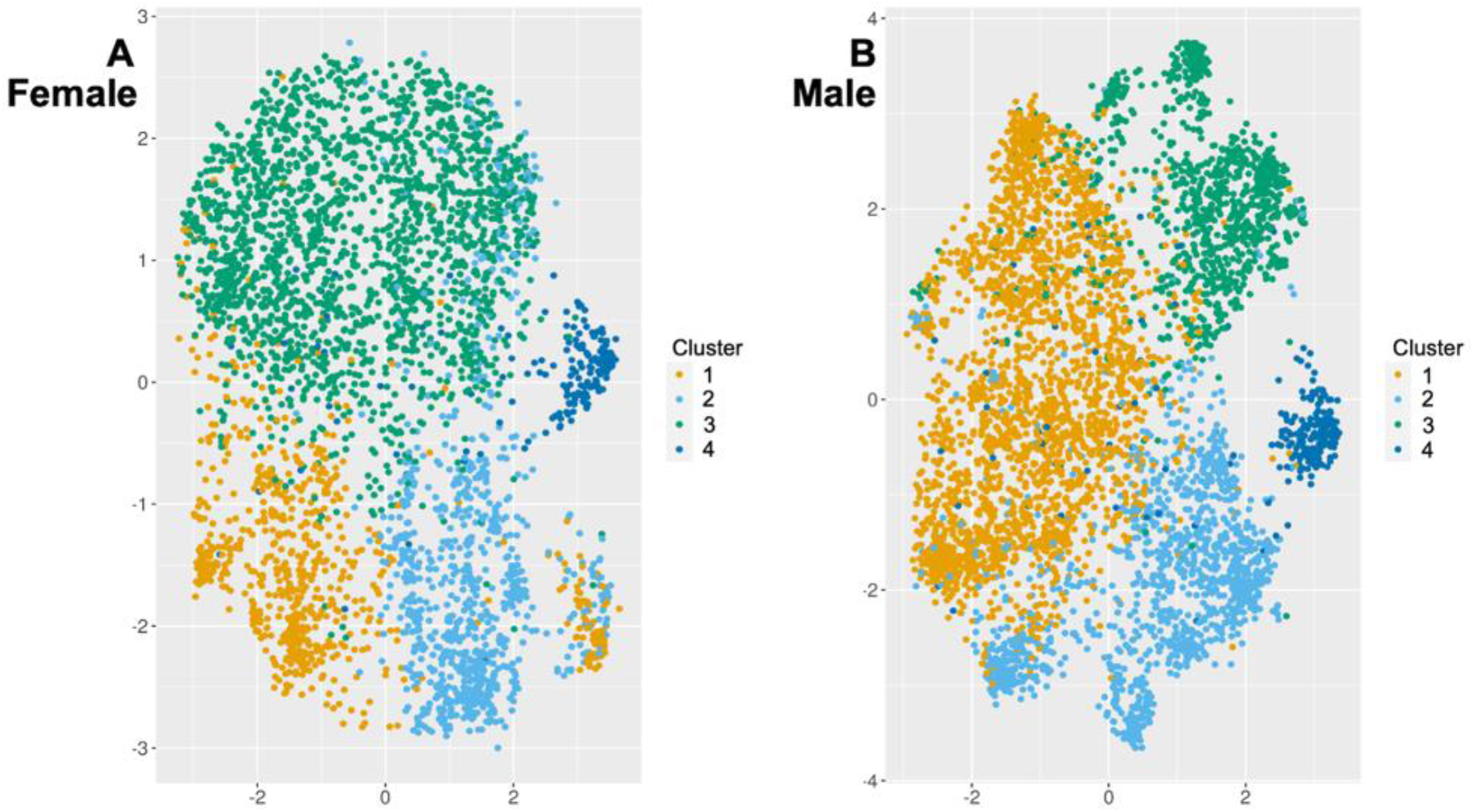
Uniform Manifold Approximation and Projection (UMAP) visualization of sex-stratified groups for unsupervised clustering. The optimal number of clusters for each sex based on Silhouette score and within-cluster and between-cluster sums of squares was four.

### Statistical Analysis of Intercluster Differences

Descriptive statistics are reported as frequencies and percentages for categorical variables and mean (standard deviation [SD]) for continuous variables. Categorical variables were compared using Pearson’s Chi-squared test or Fisher’s exact test as appropriate, and continuous variables were compared using the Wilcoxon rank sum test. All statistical testing was 2-tailed, with an α threshold of p < 0.05 used for statistical significance. All analyses were performed in R version 4.1.1 (R Foundation for Statistical Computing, Vienna, Austria), R Studio Version 1.3.1093 (RStudio, Inc., Boston, Massachusetts), and Python (version 3.9). The study protocol was approved by the Institutional Review Board of the University of Pennsylvania.

## RESULTS

Characteristics of the patients in each sex-stratified set of clusters are presented in Table 1.

**Table 1.**
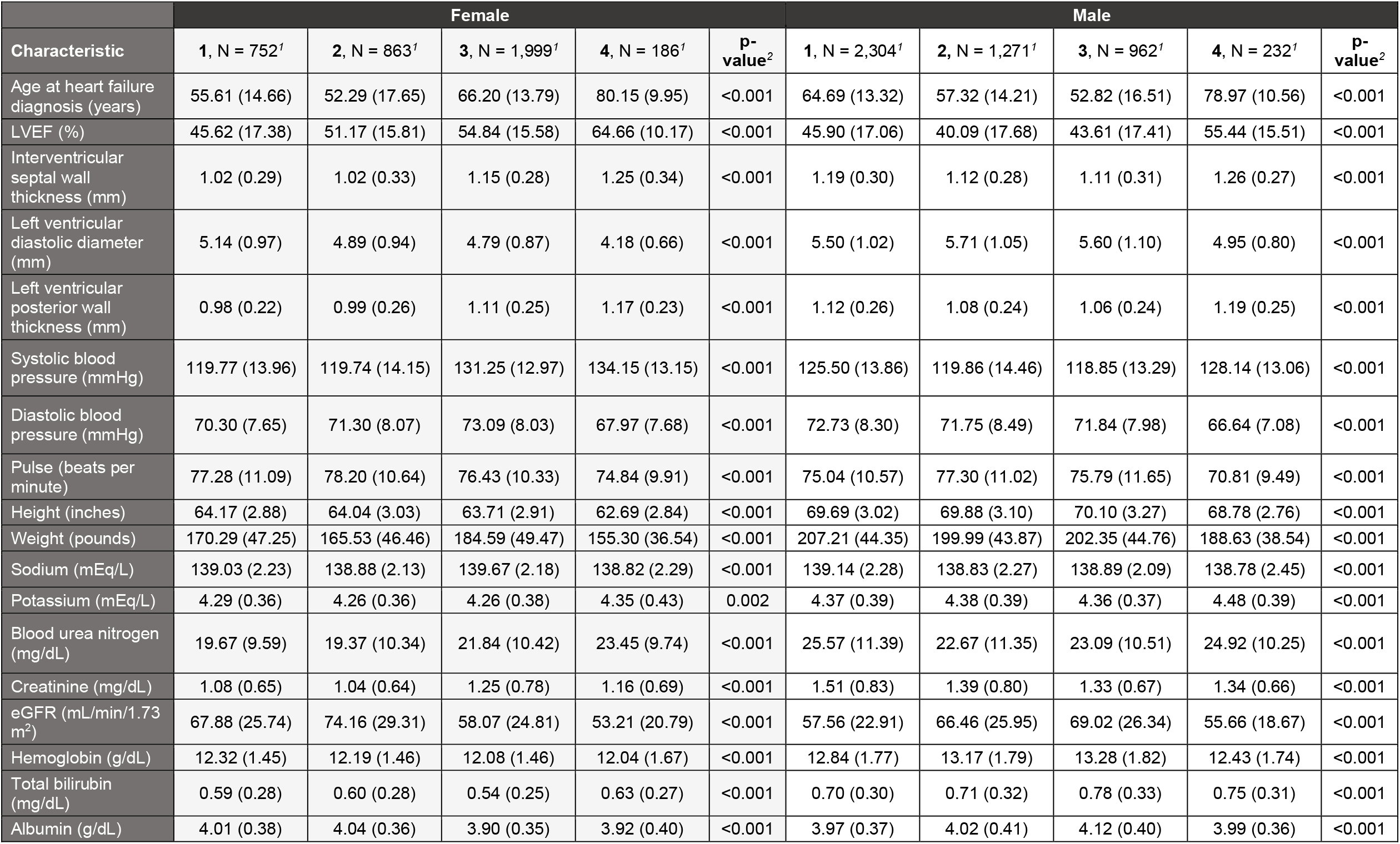
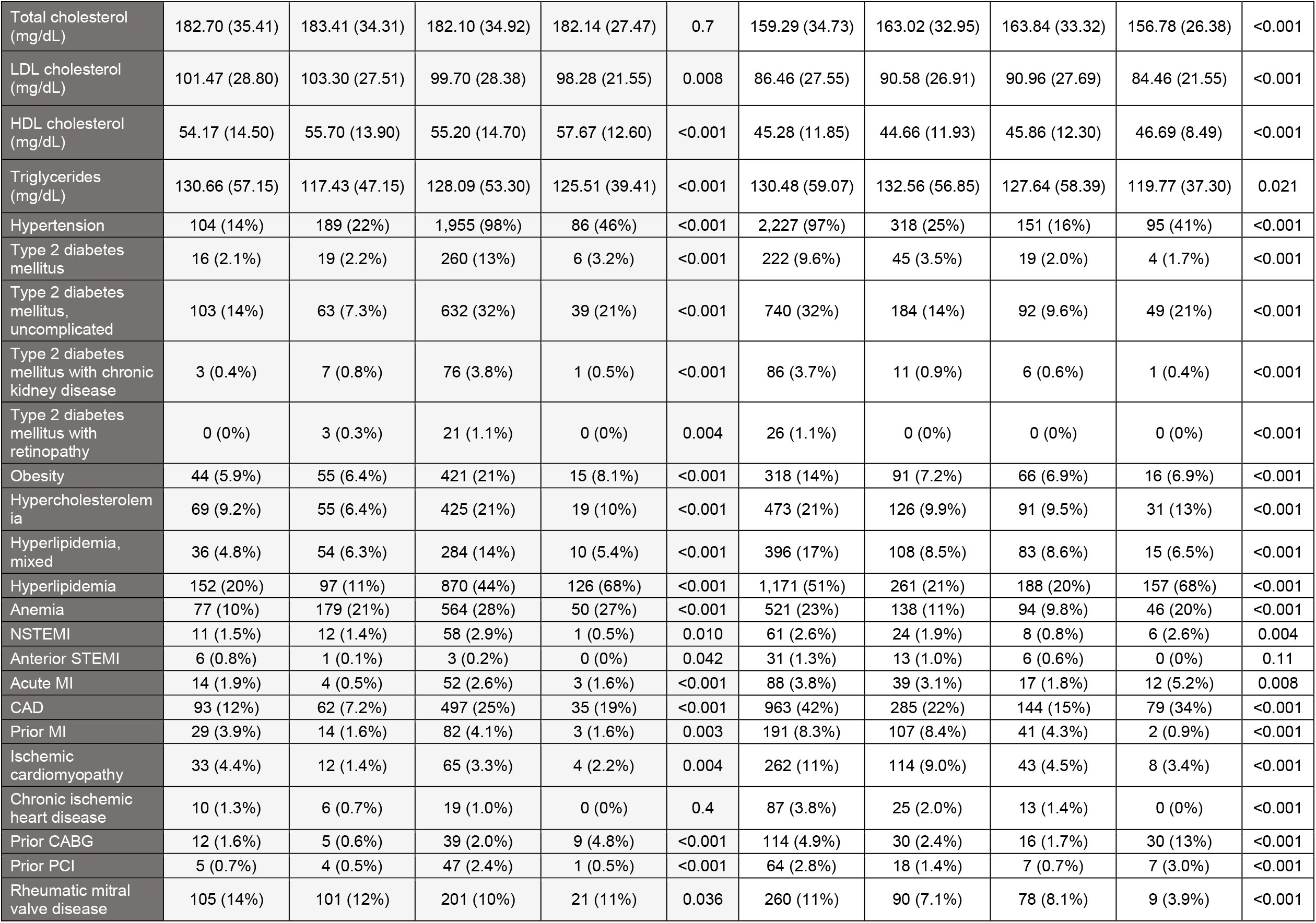
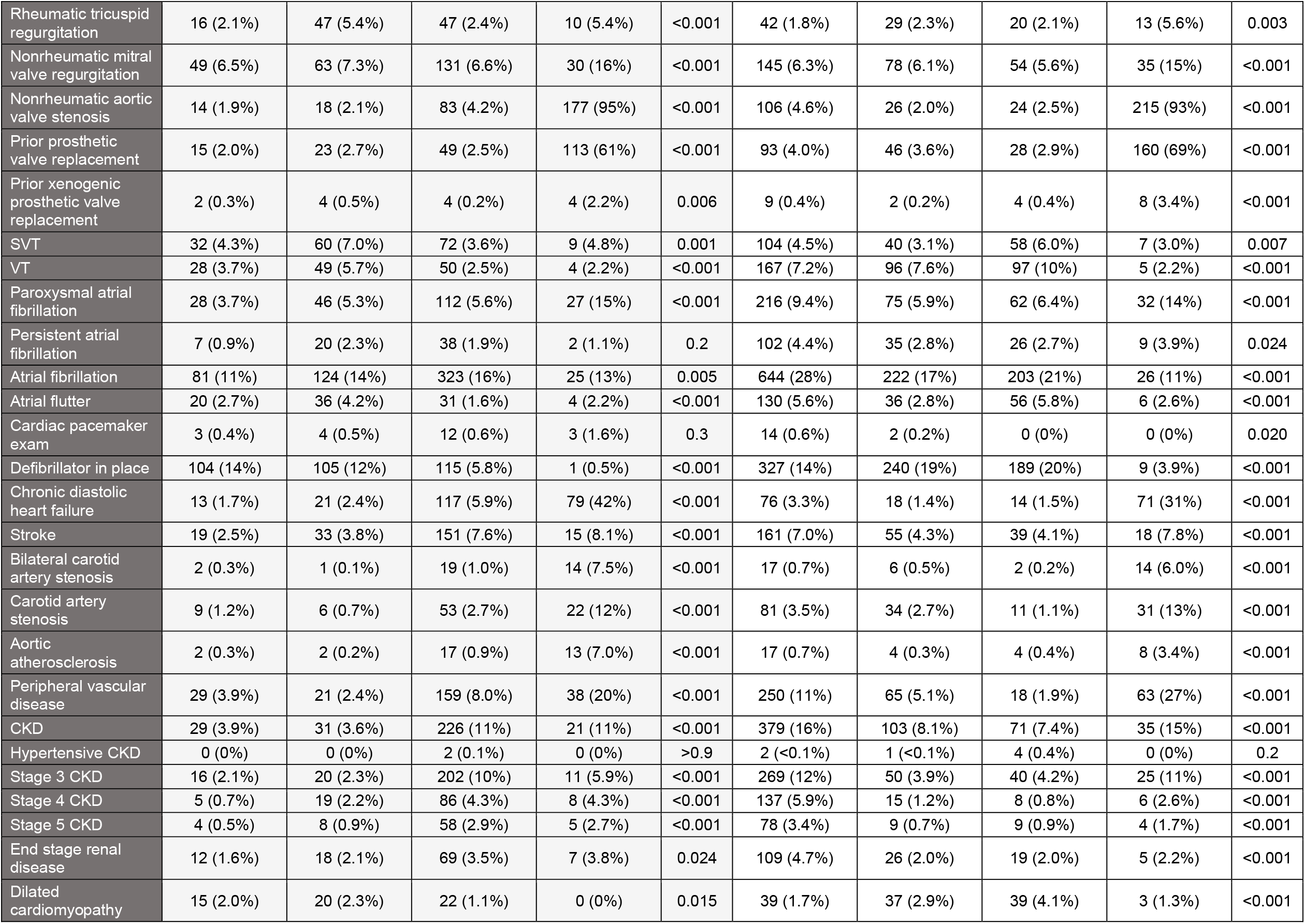
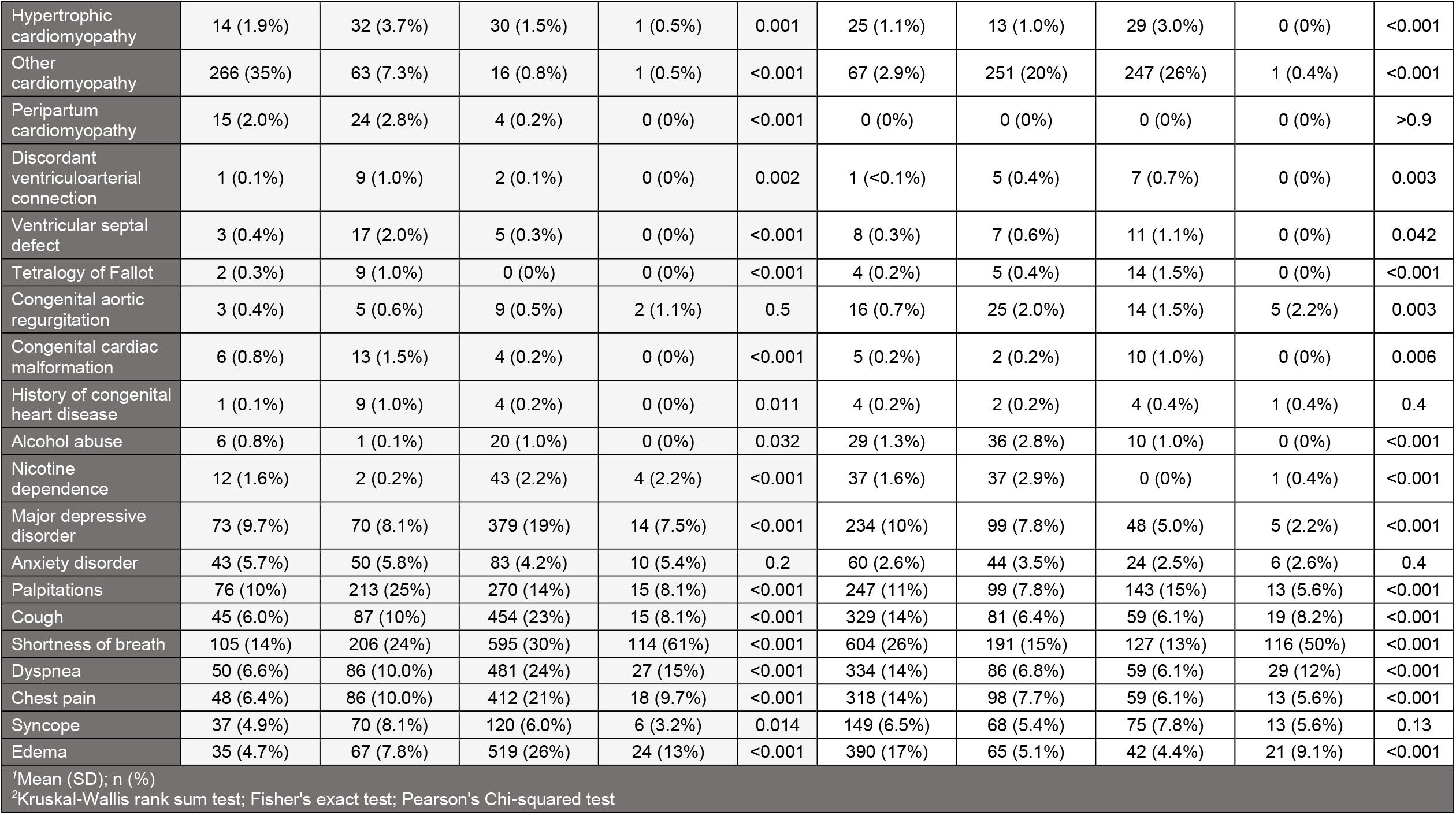
Characteristics by Patient Cluster and Stratified by Sex.

|  | Female |  |  |  |  | Male |  |  |  |  |
| --- | --- | --- | --- | --- | --- | --- | --- | --- | --- | --- |
| Characteristic | 1, N = 752<sup>1</sup> | 2, N = 863<sup>1</sup> | 3, N = 1,999<sup>1</sup> | 4, N = 186<sup>1</sup> | P-value<sup>2</sup> | 1, N = 2,304<sup>1</sup> | 2, N = 1,271<sup>1</sup> | 3, N = 962<sup>1</sup> | 4, N = 232<sup>1</sup> | P-value<sup>2</sup> |
| Age at heart failure diagnosis (years) | 55.61 (14.66) | 52.29 (17.65) | 66.20 (13.79) | 80.15 (9.95) | <0.001 | 64.69 (13.32) | 57.32 (14.21) | 52.82 (16.51) | 78.97 (10.56) | <0.001 |
| LVEF (%) | 45.62 (17.38) | 51.17 (15.81) | 54.84 (15.58) | 64.66 (10.17) | <0.001 | 45.90 (17.06) | 40.09 (17.68) | 43.61 (17.41) | 55.44 (15.51) | <0.001 |
| Interventricular septal wall thickness (mm) | 1.02 (0.29) | 1.02 (0.33) | 1.15 (0.28) | 1.25 (0.34) | <0.001 | 1.19 (0.30) | 1.12 (0.28) | 1.11 (0.31) | 1.26 (0.27) | <0.001 |
| Left ventricular diastolic diameter (mm) | 5.14 (0.97) | 4.89 (0.94) | 4.79 (0.87) | 4.18 (0.66) | <0.001 | 5.50 (1.02) | 5.71 (1.05) | 5.60 (1.10) | 4.95 (0.80) | <0.001 |
| Left ventricular posterior wall thickness (mm) | 0.98 (0.22) | 0.99 (0.26) | 1.11 (0.25) | 1.17 (0.23) | <0.001 | 1.12 (0.26) | 1.08 (0.24) | 1.06 (0.24) | 1.19 (0.25) | <0.001 |
| Systolic blood pressure (mmHg) | 119.77 (13.96) | 119.74 (14.15) | 131.25 (12.97) | 134.15 (13.15) | <0.001 | 125.50 (13.86) | 119.86 (14.46) | 118.85 (13.29) | 128.14 (13.06) | <0.001 |
| Diastolic blood pressure (mmHg) | 70.30 (7.65) | 71.30 (8.07) | 73.09 (8.03) | 67.97 (7.68) | <0.001 | 72.73 (8.30) | 71.75 (8.49) | 71.84 (7.98) | 66.64 (7.08) | <0.001 |
| Pulse (beats per minute) | 77.28 (11.09) | 78.20 (10.64) | 76.43 (10.33) | 74.84 (9.91) | <0.001 | 75.04 (10.57) | 77.30 (11.02) | 75.79 (11.65) | 70.81 (9.49) | <0.001 |
| Height (inches) | 64.17 (2.88) | 64.04 (3.03) | 63.71 (2.91) | 62.69 (2.84) | <0.001 | 69.69 (3.02) | 69.88 (3.10) | 70.10 (3.27) | 68.78 (2.76) | <0.001 |
| Weight (pounds) | 170.29 (47.25) | 165.53 (46.46) | 184.59 (49.47) | 155.30 (36.54) | <0.001 | 207.21 (44.35) | 199.99 (43.87) | 202.35 (44.76) | 188.63 (38.54) | <0.001 |
| Sodium (mEq/L) | 139.03 (2.23) | 138.88 (2.13) | 139.67 (2.18) | 138.82 (2.29) | <0.001 | 139.14 (2.28) | 138.83 (2.27) | 138.89 (2.09) | 138.78 (2.45) | <0.001 |
| Potassium (mEq/L) | 4.29 (0.36) | 4.26 (0.36) | 4.26 (0.38) | 4.35 (0.43) | 0.002 | 4.37 (0.39) | 4.38 (0.39) | 4.36 (0.37) | 4.48 (0.39) | <0.001 |
| Blood urea nitrogen (mg/dL) | 19.67 (9.59) | 19.37 (10.34) | 21.84 (10.42) | 23.45 (9.74) | <0.001 | 25.57 (11.39) | 22.67 (11.35) | 23.09 (10.51) | 24.92 (10.25) | <0.001 |
| Creatinine (mg/dL) | 1.08 (0.65) | 1.04 (0.64) | 1.25 (0.78) | 1.16 (0.69) | <0.001 | 1.51 (0.83) | 1.39 (0.80) | 1.33 (0.67) | 1.34 (0.66) | <0.001 |
| eGFR (mL/min/1.73 m<sup>2</sup>) | 67.88 (25.74) | 74.16 (29.31) | 58.07 (24.81) | 53.21 (20.79) | <0.001 | 57.56 (22.91) | 66.46 (25.95) | 69.02 (26.34) | 55.66 (18.67) | <0.001 |
| Hemoglobin (g/dL) | 12.32 (1.45) | 12.19 (1.46) | 12.08 (1.46) | 12.04 (1.67) | <0.001 | 12.84 (1.77) | 13.17 (1.79) | 13.28 (1.82) | 12.43 (1.74) | <0.001 |
| Total bilirubin (mg/dL) | 0.59 (0.28) | 0.60 (0.28) | 0.54 (0.25) | 0.63 (0.27) | <0.001 | 0.70 (0.30) | 0.71 (0.32) | 0.78 (0.33) | 0.75 (0.31) | <0.001 |
| Albumin (g/dL) | 4.01 (0.38) | 4.04 (0.36) | 3.90 (0.35) | 3.92 (0.40) | <0.001 | 3.97 (0.37) | 4.02 (0.41) | 4.12 (0.40) | 3.99 (0.36) | <0.001 |
| Total cholesterol (mg/dL) | 182.70 (35.41) | 183.41 (34.31) | 182.10 (34.92) | 182.14 (27.47) | 0.7 | 159.29 (34.73) | 163.02 (32.95) | 163.84 (33.32) | 156.78 (26.38) | <0.001 |
| LDL cholesterol (mg/dL) | 101.47 (28.80) | 103.30 (27.51) | 99.70 (28.38) | 98.28 (21.55) | 0.008 | 86.46 (27.55) | 90.58 (26.91) | 90.96 (27.69) | 84.46 (21.55) | <0.001 |
| HDL cholesterol (mg/dL) | 54.17 (14.50) | 55.70 (13.90) | 55.20 (14.70) | 57.67 (12.60) | <0.001 | 45.28 (11.85) | 44.66 (11.93) | 45.86 (12.30) | 46.69 (8.49) | <0.001 |
| Triglycerides (mg/dL) | 130.66 (57.15) | 117.43 (47.15) | 128.09 (53.30) | 125.51 (39.41) | <0.001 | 130.48 (59.07) | 132.56 (56.85) | 127.64 (58.39) | 119.77 (37.30) | 0.021 |
| Hypertension | 104 (14%) | 189 (22%) | 1,955 (98%) | 86 (46%) | <0.001 | 2,227 (97%) | 318 (25%) | 151 (16%) | 95 (41%) | <0.001 |
| Type 2 diabetes mellitus | 16 (2.1%) | 19 (2.2%) | 260 (13%) | 6 (3.2%) | <0.001 | 222 (9.6%) | 45 (3.5%) | 19 (2.0%) | 4 (1.7%) | <0.001 |
| Type 2 diabetes mellitus, uncomplicated | 103 (14%) | 63 (7.3%) | 632 (32%) | 39 (21%) | <0.001 | 740 (32%) | 184 (14%) | 92 (9.6%) | 49 (21%) | <0.001 |
| Type 2 diabetes mellitus with chronic kidney disease | 3 (0.4%) | 7 (0.8%) | 76 (3.8%) | 1 (0.5%) | <0.001 | 86 (3.7%) | 11 (0.9%) | 6 (0.6%) | 1 (0.4%) | <0.001 |
| Type 2 diabetes mellitus with retinopathy | 0 (0%) | 3 (0.3%) | 21 (1.1%) | 0 (0%) | 0.004 | 26 (1.1%) | 0 (0%) | 0 (0%) | 0 (0%) | <0.001 |
| Obesity | 44 (5.9%) | 55 (6.4%) | 421 (21%) | 15 (8.1%) | <0.001 | 318 (14%) | 91 (7.2%) | 66 (6.9%) | 16 (6.9%) | <0.001 |
| Hypercholesterolemia | 69 (9.2%) | 55 (6.4%) | 425 (21%) | 19 (10%) | <0.001 | 473 (21%) | 126 (9.9%) | 91 (9.5%) | 31 (13%) | <0.001 |
| Hyperlipidemia, mixed | 36 (4.8%) | 54 (6.3%) | 284 (14%) | 10 (5.4%) | <0.001 | 396 (17%) | 108 (8.5%) | 83 (8.6%) | 15 (6.5%) | <0.001 |
| Hyperlipidemia | 152 (20%) | 97 (11%) | 870 (44%) | 126 (68%) | <0.001 | 1,171 (51%) | 261 (21%) | 188 (20%) | 157 (68%) | <0.001 |
| Anemia | 77 (10%) | 179 (21%) | 564 (28%) | 50 (27%) | <0.001 | 521 (23%) | 138 (11%) | 94 (9.8%) | 46 (20%) | <0.001 |
| NSTEMI | 11 (1.5%) | 12 (1.4%) | 58 (2.9%) | 1 (0.5%) | 0.010 | 61 (2.6%) | 24 (1.9%) | 8 (0.8%) | 6 (2.6%) | 0.004 |
| Anterior STEMI | 6 (0.8%) | 1 (0.1%) | 3 (0.2%) | 0 (0%) | 0.042 | 31 (1.3%) | 13 (1.0%) | 6 (0.6%) | 0 (0%) | 0.11 |
| Acute MI | 14 (1.9%) | 4 (0.5%) | 52 (2.6%) | 3 (1.6%) | <0.001 | 88 (3.8%) | 39 (3.1%) | 17 (1.8%) | 12 (5.2%) | 0.008 |
| CAD | 93 (12%) | 62 (7.2%) | 497 (25%) | 35 (19%) | <0.001 | 963 (42%) | 285 (22%) | 144 (15%) | 79 (34%) | <0.001 |
| Prior MI | 29 (3.9%) | 14 (1.6%) | 82 (4.1%) | 3 (1.6%) | 0.003 | 191 (8.3%) | 107 (8.4%) | 41 (4.3%) | 2 (0.9%) | <0.001 |
| Ischemic cardiomyopathy | 33 (4.4%) | 12 (1.4%) | 65 (3.3%) | 4 (2.2%) | 0.004 | 262 (11%) | 114 (9.0%) | 43 (4.5%) | 8 (3.4%) | <0.001 |
| Chronic ischemic heart disease | 10 (1.3%) | 6 (0.7%) | 19 (1.0%) | 0 (0%) | 0.4 | 87 (3.8%) | 25 (2.0%) | 13 (1.4%) | 0 (0%) | <0.001 |
| Prior CABG | 12 (1.6%) | 5 (0.6%) | 39 (2.0%) | 9 (4.8%) | <0.001 | 114 (4.9%) | 30 (2.4%) | 16 (1.7%) | 30 (13%) | <0.001 |
| Prior PCI | 5 (0.7%) | 4 (0.5%) | 47 (2.4%) | 1 (0.5%) | <0.001 | 64 (2.8%) | 18 (1.4%) | 7 (0.7%) | 7 (3.0%) | <0.001 |
| Rheumatic mitral valve disease | 105 (14%) | 101 (12%) | 201 (10%) | 21 (11%) | 0.036 | 260 (11%) | 90 (7.1%) | 78 (8.1%) | 9 (3.9%) | <0.001 |
| Rheumatic tricuspid regurgitation | 16 (2.1%) | 47 (5.4%) | 47 (2.4%) | 10 (5.4%) | <0.001 | 42 (1.8%) | 29 (2.3%) | 20 (2.1%) | 13 (5.6%) | 0.003 |
| Nonrheumatic mitral valve regurgitation | 49 (6.5%) | 63 (7.3%) | 131 (6.6%) | 30 (16%) | <0.001 | 145 (6.3%) | 78 (6.1%) | 54 (5.6%) | 35 (15%) | <0.001 |
| Nonrheumatic aortic valve stenosis | 14 (1.9%) | 18 (2.1%) | 83 (4.2%) | 177 (95%) | <0.001 | 106 (4.6%) | 26 (2.0%) | 24 (2.5%) | 215 (93%) | <0.001 |
| Prior prosthetic valve replacement | 15 (2.0%) | 23 (2.7%) | 49 (2.5%) | 113 (61%) | <0.001 | 93 (4.0%) | 46 (3.6%) | 28 (2.9%) | 160 (69%) | <0.001 |
| Prior xenogenic prosthetic valve replacement | 2 (0.3%) | 4 (0.5%) | 4 (0.2%) | 4 (2.2%) | 0.006 | 9 (0.4%) | 2 (0.2%) | 4 (0.4%) | 8 (3.4%) | <0.001 |
| SVT | 32 (4.3%) | 60 (7.0%) | 72 (3.6%) | 9 (4.8%) | 0.001 | 104 (4.5%) | 40 (3.1%) | 58 (6.0%) | 7 (3.0%) | 0.007 |
| VT | 28 (3.7%) | 49 (5.7%) | 50 (2.5%) | 4 (2.2%) | <0.001 | 167 (7.2%) | 96 (7.6%) | 97 (10%) | 5 (2.2%) | <0.001 |
| Paroxysmal atrial fibrillation | 28 (3.7%) | 46 (5.3%) | 112 (5.6%) | 27 (15%) | <0.001 | 216 (9.4%) | 75 (5.9%) | 62 (6.4%) | 32 (14%) | <0.001 |
| Persistent atrial fibrillation | 7 (0.9%) | 20 (2.3%) | 38 (1.9%) | 2 (1.1%) | 0.2 | 102 (4.4%) | 35 (2.8%) | 26 (2.7%) | 9 (3.9%) | 0.024 |
| Atrial fibrillation | 81 (11%) | 124 (14%) | 323 (16%) | 25 (13%) | 0.005 | 644 (28%) | 222 (17%) | 203 (21%) | 26 (11%) | <0.001 |
| Atrial flutter | 20 (2.7%) | 36 (4.2%) | 31 (1.6%) | 4 (2.2%) | <0.001 | 130 (5.6%) | 36 (2.8%) | 56 (5.8%) | 6 (2.6%) | <0.001 |
| Cardiac pacemaker exam | 3 (0.4%) | 4 (0.5%) | 12 (0.6%) | 3 (1.6%) | 0.3 | 14 (0.6%) | 2 (0.2%) | 0 (0%) | 0 (0%) | 0.020 |
| Defibrillator in place | 104 (14%) | 105 (12%) | 115 (5.8%) | 1 (0.5%) | <0.001 | 327 (14%) | 240 (19%) | 189 (20%) | 9 (3.9%) | <0.001 |
| Chronic diastolic heart failure | 13 (1.7%) | 21 (2.4%) | 117 (5.9%) | 79 (42%) | <0.001 | 76 (3.3%) | 18 (1.4%) | 14 (1.5%) | 71 (31%) | <0.001 |
| Stroke | 19 (2.5%) | 33 (3.8%) | 151 (7.6%) | 15 (8.1%) | <0.001 | 161 (7.0%) | 55 (4.3%) | 39 (4.1%) | 18 (7.8%) | <0.001 |
| Bilateral carotid artery stenosis | 2 (0.3%) | 1 (0.1%) | 19 (1.0%) | 14 (7.5%) | <0.001 | 17 (0.7%) | 6 (0.5%) | 2 (0.2%) | 14 (6.0%) | <0.001 |
| Carotid artery stenosis | 9 (1.2%) | 6 (0.7%) | 53 (2.7%) | 22 (12%) | <0.001 | 81 (3.5%) | 34 (2.7%) | 11 (1.1%) | 31 (13%) | <0.001 |
| Aortic atherosclerosis | 2 (0.3%) | 2 (0.2%) | 17 (0.9%) | 13 (7.0%) | <0.001 | 17 (0.7%) | 4 (0.3%) | 4 (0.4%) | 8 (3.4%) | <0.001 |
| Peripheral vascular disease | 29 (3.9%) | 21 (2.4%) | 159 (8.0%) | 38 (20%) | <0.001 | 250 (11%) | 65 (5.1%) | 18 (1.9%) | 63 (27%) | <0.001 |
| CKD | 29 (3.9%) | 31 (3.6%) | 226 (11%) | 21 (11%) | <0.001 | 379 (16%) | 103 (8.1%) | 71 (7.4%) | 35 (15%) | <0.001 |
| Hypertensive CKD | 0 (0%) | 0 (0%) | 2 (0.1%) | 0 (0%) | >0.9 | 2 (<0.1%) | 1 (<0.1%) | 4 (0.4%) | 0 (0%) | 0.2 |
| Stage 3 CKD | 16 (2.1%) | 20 (2.3%) | 202 (10%) | 11 (5.9%) | <0.001 | 269 (12%) | 50 (3.9%) | 40 (4.2%) | 25 (11%) | <0.001 |
| Stage 4 CKD | 5 (0.7%) | 19 (2.2%) | 86 (4.3%) | 8 (4.3%) | <0.001 | 137 (5.9%) | 15 (1.2%) | 8 (0.8%) | 6 (2.6%) | <0.001 |
| Stage 5 CKD | 4 (0.5%) | 8 (0.9%) | 58 (2.9%) | 5 (2.7%) | <0.001 | 78 (3.4%) | 9 (0.7%) | 9 (0.9%) | 4 (1.7%) | <0.001 |
| End stage renal disease | 12 (1.6%) | 18 (2.1%) | 69 (3.5%) | 7 (3.8%) | 0.024 | 109 (4.7%) | 26 (2.0%) | 19 (2.0%) | 5 (2.2%) | <0.001 |
| Dilated cardiomyopathy | 15 (2.0%) | 20 (2.3%) | 22 (1.1%) | 0 (0%) | 0.015 | 39 (1.7%) | 37 (2.9%) | 39 (4.1%) | 3 (1.3%) | <0.001 |
| Hypertrophic cardiomyopathy | 14 (1.9%) | 32 (3.7%) | 30 (1.5%) | 1 (0.5%) | 0.001 | 25 (1.1%) | 13 (1.0%) | 29 (3.0%) | 0 (0%) | <0.001 |
| Other cardiomyopathy | 266 (35%) | 63 (7.3%) | 16 (0.8%) | 1 (0.5%) | <0.001 | 67 (2.9%) | 251 (20%) | 247 (26%) | 1 (0.4%) | <0.001 |
| Peripartum cardiomyopathy | 15 (2.0%) | 24 (2.8%) | 4 (0.2%) | 0 (0%) | <0.001 | 0 (0%) | 0 (0%) | 0 (0%) | 0 (0%) | >0.9 |
| Discordant ventriculoarterial connection | 1 (0.1%) | 9 (1.0%) | 2 (0.1%) | 0 (0%) | 0.002 | 1 (<0.1%) | 5 (0.4%) | 7 (0.7%) | 0 (0%) | 0.003 |
| Ventricular septal defect | 3 (0.4%) | 17 (2.0%) | 5 (0.3%) | 0 (0%) | <0.001 | 8 (0.3%) | 7 (0.6%) | 11 (1.1%) | 0 (0%) | 0.042 |
| Tetralogy of Fallot | 2 (0.3%) | 9 (1.0%) | 0 (0%) | 0 (0%) | <0.001 | 4 (0.2%) | 5 (0.4%) | 14 (1.5%) | 0 (0%) | <0.001 |
| Congenital aortic regurgitation | 3 (0.4%) | 5 (0.6%) | 9 (0.5%) | 2 (1.1%) | 0.5 | 16 (0.7%) | 25 (2.0%) | 14 (1.5%) | 5 (2.2%) | 0.003 |
| Congenital cardiac malformation | 6 (0.8%) | 13 (1.5%) | 4 (0.2%) | 0 (0%) | <0.001 | 5 (0.2%) | 2 (0.2%) | 10 (1.0%) | 0 (0%) | 0.006 |
| History of congenital heart disease | 1 (0.1%) | 9 (1.0%) | 4 (0.2%) | 0 (0%) | 0.011 | 4 (0.2%) | 2 (0.2%) | 4 (0.4%) | 1 (0.4%) | 0.4 |
| Alcohol abuse | 6 (0.8%) | 1 (0.1%) | 20 (1.0%) | 0 (0%) | 0.032 | 29 (1.3%) | 36 (2.8%) | 10 (1.0%) | 0 (0%) | <0.001 |
| Nicotine dependence | 12 (1.6%) | 2 (0.2%) | 43 (2.2%) | 4 (2.2%) | <0.001 | 37 (1.6%) | 37 (2.9%) | 0 (0%) | 1 (0.4%) | <0.001 |
| Major depressive disorder | 73 (9.7%) | 70 (8.1%) | 379 (19%) | 14 (7.5%) | <0.001 | 234 (10%) | 99 (7.8%) | 48 (5.0%) | 5 (2.2%) | <0.001 |
| Anxiety disorder | 43 (5.7%) | 50 (5.8%) | 83 (4.2%) | 10 (5.4%) | 0.2 | 60 (2.6%) | 44 (3.5%) | 24 (2.5%) | 6 (2.6%) | 0.4 |
| Palpitations | 76 (10%) | 213 (25%) | 270 (14%) | 15 (8.1%) | <0.001 | 247 (11%) | 99 (7.8%) | 143 (15%) | 13 (5.6%) | <0.001 |
| Cough | 45 (6.0%) | 87 (10%) | 454 (23%) | 15 (8.1%) | <0.001 | 329 (14%) | 81 (6.4%) | 59 (6.1%) | 19 (8.2%) | <0.001 |
| Shortness of breath | 105 (14%) | 206 (24%) | 595 (30%) | 114 (61%) | <0.001 | 604 (26%) | 191 (15%) | 127 (13%) | 116 (50%) | <0.001 |
| Dyspnea | 50 (6.6%) | 86 (10.0%) | 481 (24%) | 27 (15%) | <0.001 | 334 (14%) | 86 (6.8%) | 59 (6.1%) | 29 (12%) | <0.001 |
| Chest pain | 48 (6.4%) | 86 (10.0%) | 412 (21%) | 18 (9.7%) | <0.001 | 318 (14%) | 98 (7.7%) | 59 (6.1%) | 13 (5.6%) | <0.001 |
| Syncope | 37 (4.9%) | 70 (8.1%) | 120 (6.0%) | 6 (3.2%) | 0.014 | 149 (6.5%) | 68 (5.4%) | 75 (7.8%) | 13 (5.6%) | 0.13 |
| Edema | 35 (4.7%) | 67 (7.8%) | 519 (26%) | 24 (13%) | <0.001 | 390 (17%) | 65 (5.1%) | 42 (4.4%) | 21 (9.1%) | <0.001 |

<sup>1</sup>Mean (SD); n (%)
<sup>2</sup>Kruskal-Wallis rank sum test; Fisher's exact test; Pearson's Chi-squared test

### Female-stratified Clusters

Patients in Cluster 1 of 4 (N=752; 19.8%) were more likely to have a non-ischemic cardiomyopathy and the largest left ventricular end diastolic diameters (5.14 millimeters ± 0.97). Most individuals with an implantable cardioverter defibrillator in place (14%) were grouped into this cluster (Figure 5A). Current (14%) or past tobacco use (86%) was highly prevalent. Cluster 2 of 4 (N=863; 22.7%) was comprised of females who were diagnosed with HF at the youngest age (mean 52.3 years ± 17.7) and most commonly had palpitations (25%). Myocardial structural abnormalities, including atrial and ventricular septal defects, Tetralogy of Fallot, and discordant ventriculoarterial connection, defined this cluster. These individuals had the highest albumin and estimated glomerular filtration rates (Figure 5B). Cluster 3 of 4 (N=1999; 52.6%) was characterized by a high prevalence of hypertension (98%), type 2 diabetes (32%), hyperlipidemia (44%), and obesity. These patients’ symptom burden most frequently included edema (26%), dyspnea, and cough. Assignment into this cluster was also driven by traits that indicated higher rates of health maintenance activities, i.e., immunization visits and age-appropriate cancer screenings (Figure 5C). Females in Cluster 4 of 4 (N=186; 4.9%) were the oldest (mean age 80.2 years ± 10) with the highest prevalence of aortic stenosis (95%) and prosthetic valve replacement (61%), diastolic heart failure (42%), and atherosclerotic cardiovascular disease as demonstrated by the presence of carotid artery stenosis, aortic atherosclerosis, and peripheral vascular disease (Figure 5D).

**FIGURE 5.**
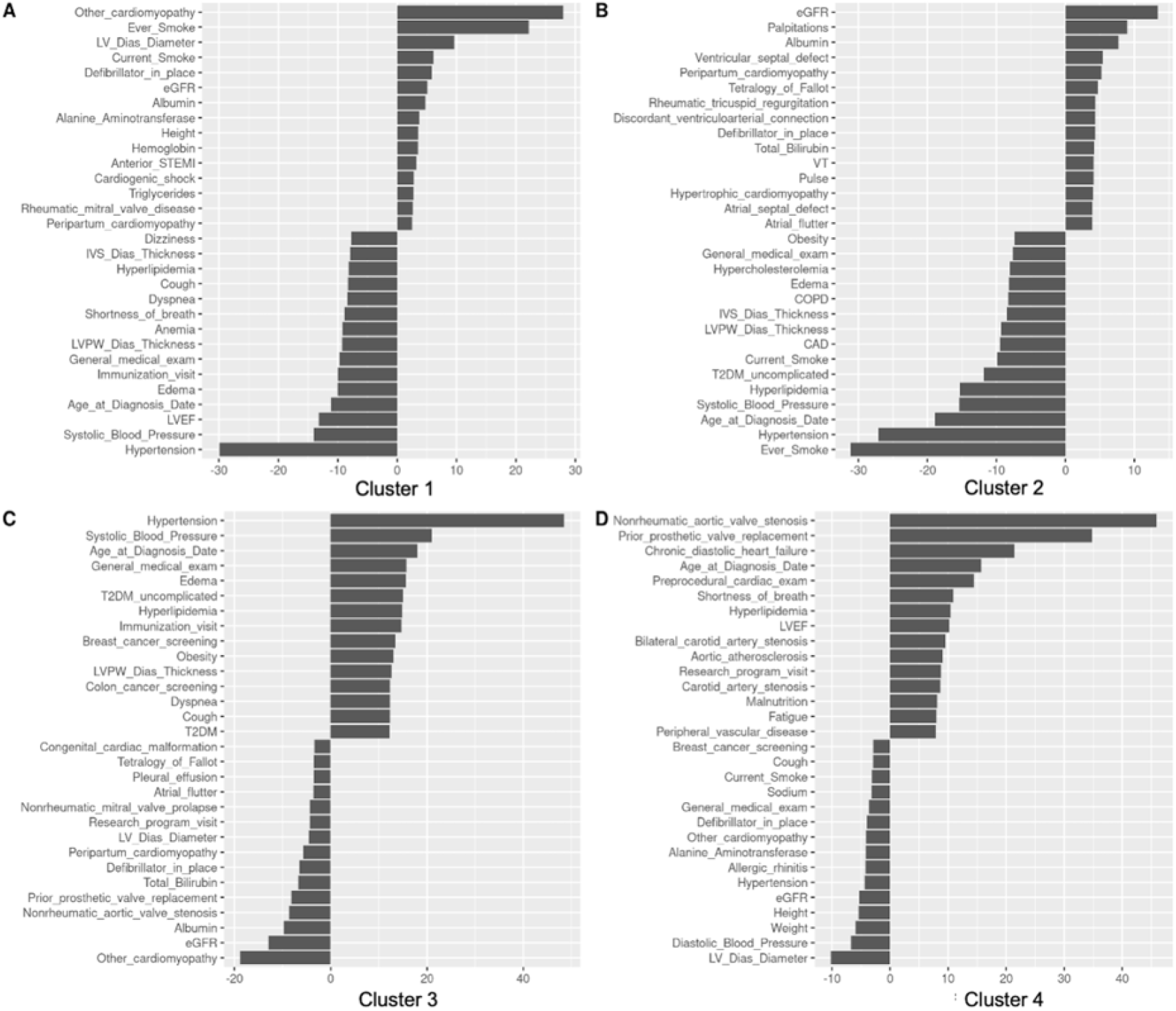
Visual representation of test values comparing the proportion of electronic health record-derived input features (y-axis) within each female-only cluster group (1-4) with the proportion of the population individuals with each input feature in the female-only population. Positive test values indicate higher prevalence or higher values of the input feature; negative test values indicate lower prevalence or lower values the input feature.

### Male-stratified Clusters

Males with cardiometabolic comorbidities, including hypertension (97%), hyperlipidemia (51%), coronary artery disease (42%), and type 2 diabetes (32%), were largely assigned to Cluster 1 (N=2304; 48.3%) (Figure 6A). Cluster 2 (N=1271, 26.7%) was characterized by higher relative prevalences of tobacco and alcohol use and by individuals who more often had dilated cardiomyopathies (Figure 6B). Cluster 3 (N=962; 20.2%) was disproportionately composed of males with congenital or inherited heart diseases such as tetralogy of Fallot, hypertrophic cardiomyopathy, and other congenital cardiac malformations. Hepatic and renal function were of importance in this cluster (Figure 6C). Similar to the females, the males in Cluster 4 (N=232; 4.8%) more often had aortic valve stenosis (93%), prosthetic heart valves (69%), diastolic heart failure (31%), and atherosclerotic cardiovascular disease manifested by peripheral vascular disease, carotid artery disease, and prior coronary artery bypass surgery (Figure 6D).

**FIGURE 6.**
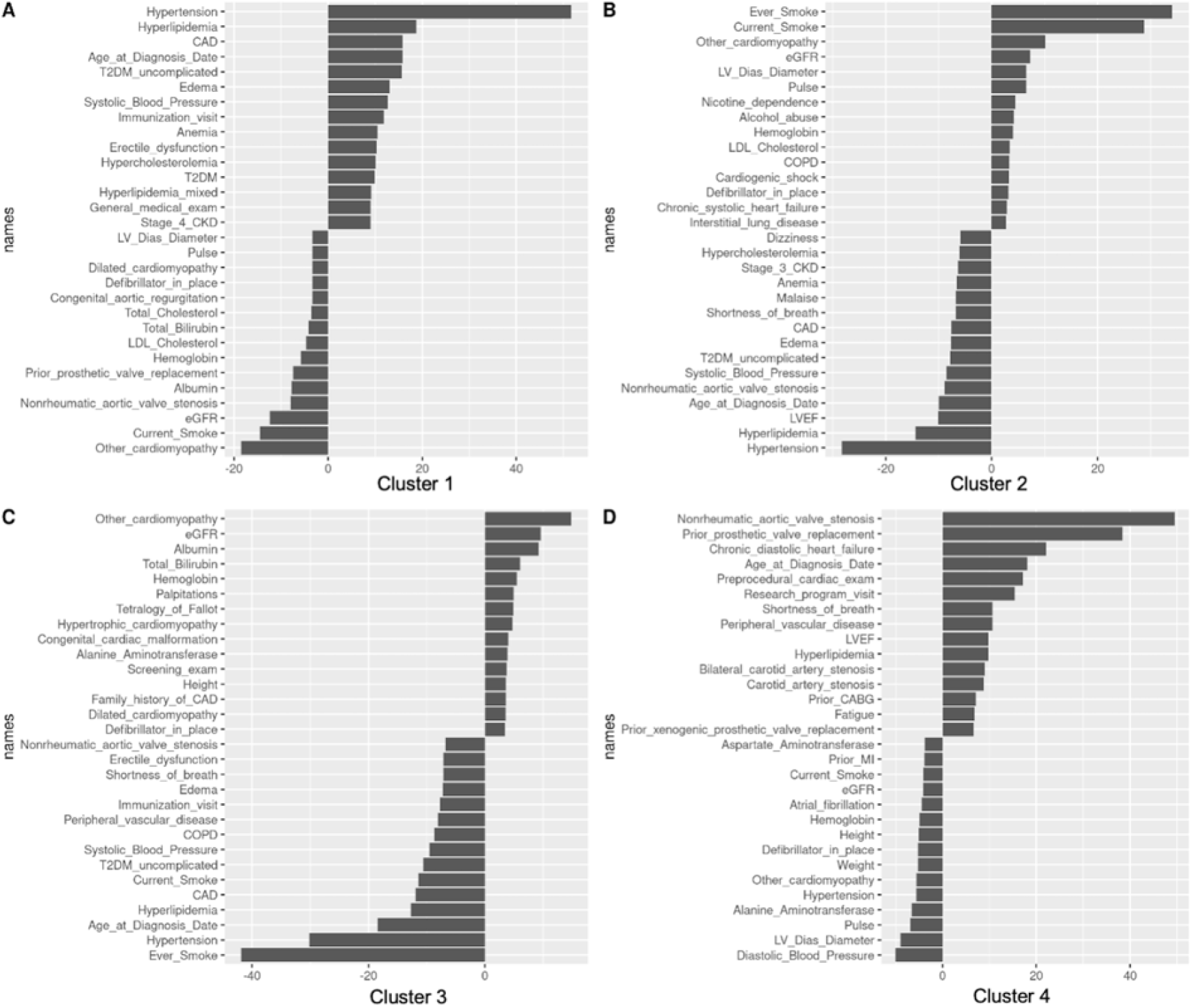
Visual representation of test values comparing the proportion of electronic health record-derived input features (y-axis) within each male-only cluster group (1-4) with the proportion of the population individuals with each input feature in the male-only population. Positive test values indicate higher prevalence of higher values of the input feature; negative test values indicate lower prevalence or lower values of the input feature.

We united the four female and four male clusters into a single projection to visualize the similarities in cluster phenotype across both sexes (Figure 7). In both sexes, there was a cluster (female cluster 3 and male cluster 1) show significant overlap driven by cardiometabolic traits (i.e., hypertension, hyperlipidemia, obesity) and a separate cluster (female cluster 4 and male cluster 4) characterized by advanced age, aortic valve disease and valve replacement, and atherosclerotic cardiovascular disease, which are represented by a high degree of localized overlap in Figure 7.

**FIGURE 7.**
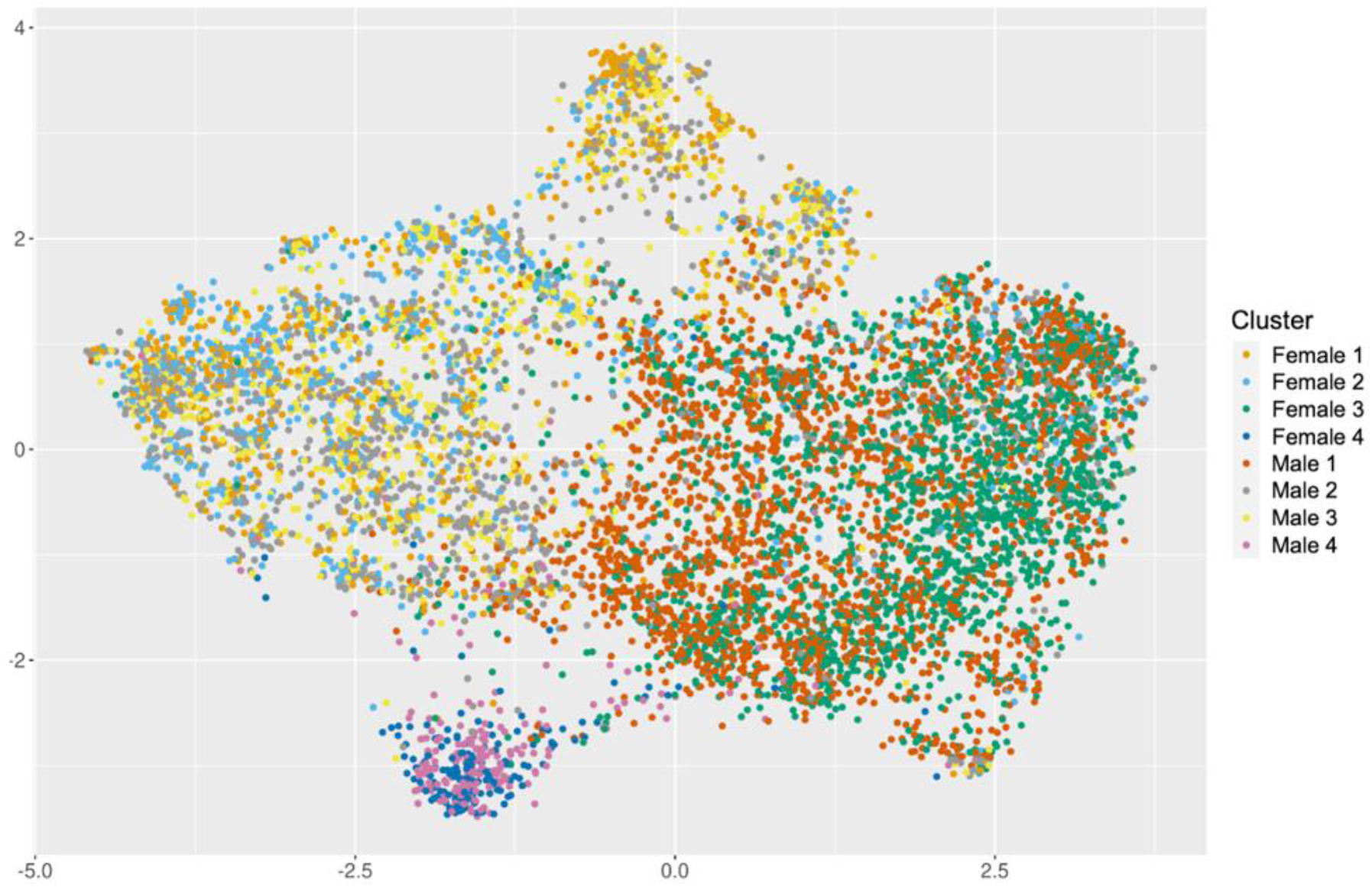
Patients with heart failure in the electronic health record-derived cohort labeled by sex and cluster assignment. Female cluster 3 and male cluster 1 and female cluster 4 and male cluster 4 show significant overlap, indicating shared phenotypic composition.

## DISCUSSION

In this proof-of-concept study, we demonstrated that an unsupervised machine learning approach applied to routine clinical data extracted from the EHR is able to define pathophysiologically relevant clusters of patients with HF. We created a bioinformatics pipeline employing PCA, UMAP, and spectral clustering and using traits routinely collected during the clinical care of patients with HF, and with this method, demonstrated that distinct subgroups of patients with HF can be identified without a priori labeling. After stratifying by sex, we showed that that each sex-specific cluster of patients had significantly different myocardial abnormalities, comorbid conditions, and symptoms compared with one another. Prior studies of cluster analysis applied to patients with HF have largely been limited to either LVEF-defined groups^2,10–13^, clinical characteristics^14–17^, or datasets derived from prospective registries^18,19^, clinical trials^6,20–23^, or administrative claims data^24^. To our knowledge, ours is the first study to deploy unsupervised learning algorithms on a broad population of patients with HF derived from the EHR.

There are known sex differences in the epidemiology, risk factors, management, treatment, and outcomes of HF^25^, and females are underrepresented in HF research.^26^ Our cohort included 44% self-identified females, a higher proportion of females than most HF clinical trials and cohort studies to date. Our strategy of sex stratification prior to cluster analysis afforded the opportunity to uncover latent characteristics of the HF phenotype that may have otherwise been masked. Interestingly, three of the four clusters were similar across both sexes and aligned with broader cardiometabolic, valvular, and structural cardiomyopathic phenotypes. This aligns with previous epidemiological research^27^ highlighting that preventive efforts targeted toward reducing the population burden of hypertension, hyperlipidemia, obesity, and diabetes still offers the greatest potential to reduce the risk of progression to HF in both females and males. Machine learning-based models integrated into the EHR could enable prospective identification of patients whose comorbidity profiles elevate the risk for incident HF and facilitate testing of sex-specific implementation approaches to primordial and primary HF prevention.

Longitudinal cohort studies have provided the foundational data on the associations between singular HF risk factors, e.g., hypertension, and incident HF and HF outcomes. However, what is less known from these types of studies are the impacts of groups of co-occurring comorbid diseases or risk factors. With this unsupervised learning approach, we are more readily able to identify specific patterns or clusters of comorbidities that together may influence an individual’s HF clinical trajectory differently compared with other patterns. While we were unable to correlate these cluster assignments to outcomes in this study, future analyses evaluating these associations between cluster profiles and cardiovascular outcomes, including healthcare utilization, are planned to further explore these hypotheses.

Notably, LVEF was largely absent from the input features most responsible for determining cluster assignment in our analysis. This has been previously demonstrated in an unsupervised cluster analysis applied to an HF administrative claims dataset in which none of the derived clusters tracked with HF subtype defined by LVEF.^24^ The limitations of LVEF in categorizing HF are well-recognized^28^, and several therapies have been proven to be of benefit regardless of LVEF.^29,30^ Recent changes in the nomenclature of HF subtype classification^31^ have signaled a reconsideration of the determinants of patient eligibility for HF therapies. The results of our cluster analysis align with this position by demonstrating the importance of the underlying pathophysiological features of HF. Additional uses of this EHR-based unsupervised cluster analysis include identification of patients who might derive greatest benefit from specific therapies, e.g., sodium-glucose cotransporter-2 inhibitors for patients in the cardiometabolic HF clusters, and targeted enrollment of patients into HF clinical trials across the range of LVEF.

Limitations of our study should be noted. Input features were restricted to those available in our EHR data warehouse, and our study only included patients from one health system, which may result in selection bias. Other biologically measured features, such as electrocardiographic tracings that have been shown to have predictive value for disease state classification and prognosis^32–34^, were intentionally not incorporated into this analysis to test the scalability of our approach using only EHR-derived traits. Comorbidity and symptom profiles were derived from presence of ICD-10-CM codes, and misclassification is possible. Future unsupervised learning analyses incorporating data generated by patients are an opportunity for further refinement of this concept.

In conclusion, readily available EHR data collected in the course of routine care can be leveraged to classify patients into major phenotypic HF subtypes using data-driven approaches. Our analysis highlights the impact of sex on HF subtypes and the limited impact of LVEF compared to other pathophysiologic input features. Future work is required to refine and validate subtypes to enable assessment of prognosis and response to targeted treatment strategies.

## Data Availability

All data produced in the present study are available upon reasonable request to the authors

## Non-standard Abbreviations and Acronyms

HER: electronic health record
ICD-10-CM: International Classification of Diseases, Tenth Revision, Clinical Modification
HF: heart failure
HfpEF: heart failure with preserved ejection fraction
HfrEF: heart failure with reduced ejection fraction
LVEF: left ventricular ejection fraction
NYHA: New York Heart Association
PCA: Principal Component Analysis
UMAP: Uniform Manifold Approximation and Projection

## Acknowledgments

None.

